# L18F substrain of SARS-CoV-2 VOC-202012/01 is rapidly spreading in England

**DOI:** 10.1101/2021.02.07.21251262

**Authors:** Frederic Grabowski, Marek Kochańczyk, Tomasz Lipniacki

## Abstract

The Variant of Concern (VOC)-202012/01 (also known as B.1.1.7) is a rapidly growing lineage of SARS-CoV-2. In January 2021, VOC-202012/01 constituted about 80% of SARS-CoV-2 genomes sequenced in England and was present in 27 out of 29 countries that reported at least 50 viral genomes. As this strain will likely spread globally towards fixation, it is important to monitor its molecular evolution. Based on GISAID data we systematically estimated growth rates of mutations acquired by the VOC lineage to find that L18F substitution in viral spike protein has initiated a substrain characterized by replicative advantage of 1.70 [95% CI: 1.56–1.96] in relation to the remaining VOC-202012/01 substrains. The L18F mutation is of significance because when recently analyzed in the context of the South African strain 501Y.V2 it has been found to compromise binding of neutralizing antibodies. We additionally indicate three mutations that were acquired by VOC-202012/01 in the receptor binding motif of spike, specifically E484K, F490S, and S494P, that may also give rise to escape mutants. Such mutants may hinder efficiency of existing vaccines and expand in response to the increasing after-infection or vaccine-induced seroprevalence.

## Introduction

The SARS-CoV-2 Variant of Concern (VOC)-202012/01, also known as B.1.1.7 lineage (first GISAID sequence accession ID: EPI_ISL_601443) is characterized by nine spike protein mutations (three deletions: 69H–70V, 145V; and six substitutions: N501Y, A570D, D614G, P681H, T716I, S982A, D1118H). The lineage has started spreading rapidly in mid-October 2020 to constitute about 80% of all SARS-CoV-2 genomes sequenced in England in January 2021.^1, 2^ Its spreading capacity may be expressed in terms of its replicative advantage—the ratio of the reproduction numbers of the VOC to the non-VOC strains, 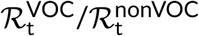. The replicative advantage of the VOC has been estimated by several groups as 1.47 [95% CI: 1.34–1.59],^3^ 1.56 [95% CI: 1.50-1.74],^4^ 1.75 [95% CrI: 1.70–1.80]^5^ or 2.24 [95% CI: 2.03–2.48].^6^ Based on GISAID data available on February 5, 2021, aggregated over whole England, using the same method as previously,^6^ we may estimate that 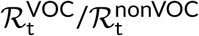 lies in the range 1.88–2.28, where the bounds are calculated correspondingly as the estimate within the time span of weeks 43–51 of 2020 (lower bound) and the estimate within the time span of weeks 43–47 of 2020 (upper bound). In such 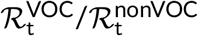 estimation it is assumed that both the VOC and the non-VOC strains have the same average serial interval of 6.73 days^7^; then, 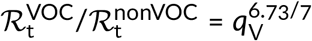, where *q*_V_is the weekly growth of the ratio of the VOC to the non-VOC strains, *r*_V_ (see Figure 1).

**Figure 1:**
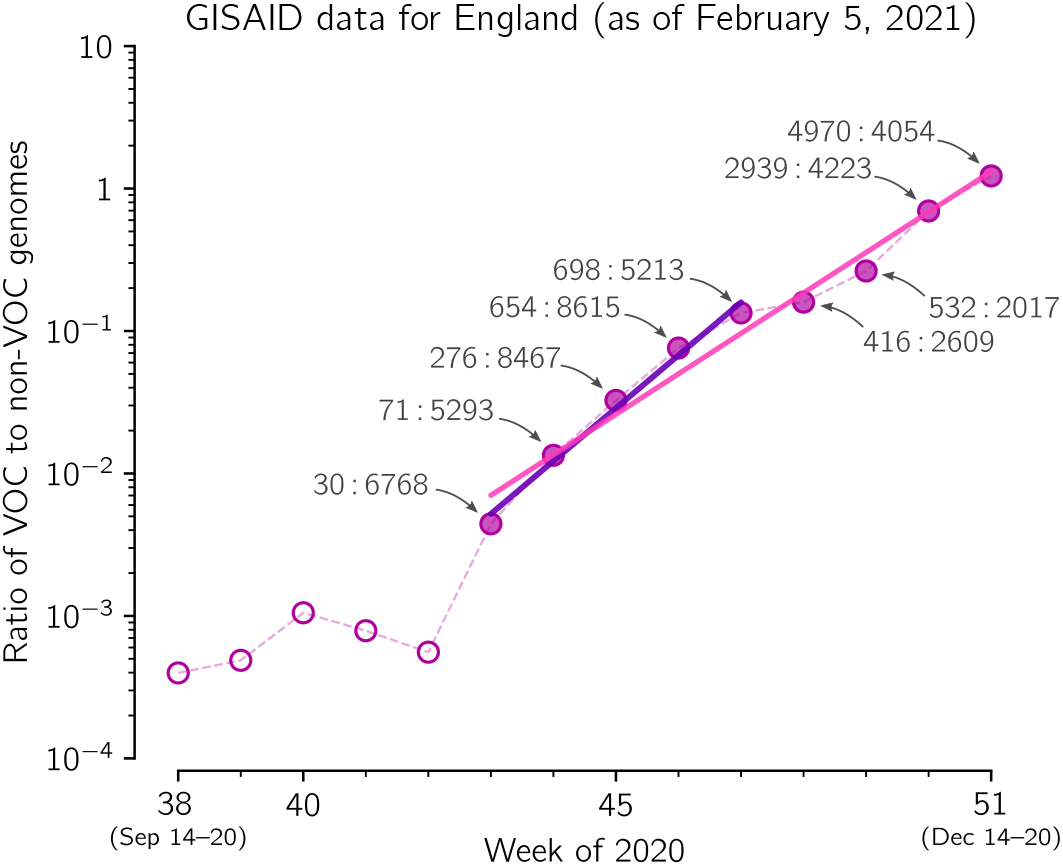
The replicative advantage of the VOC strain. The ratio of the VOC to the non-VOC genomes collected in weeks 38–51 of 2020 in England, *r*_V_. Data points are labeled with ratios of the counts of genome sequences. Data aggregated into weeks indicated that *r*_V_ changes from 30 : 6738 = 0.0045 in week 43 to 4970 : 4054 = 1.23 in week 51, i.e., 275 times, which means that *r*_VOC_ increases 275^1/8^ ≈ 2.02-fold per week. The trend line is fitted to data points for weeks 43–51 (violet) and for weeks 43–47 (pink). The weekly growth rate is 1.92 [95% CI: 1.78–2.08] for weeks 43–51 and 2.35 [95% CI: 2.10–2.62] for weeks 43–47 of 2020. The figure is based on GISAID data submitted till February 5, 2021. The VOC genomes were counted by selecting the B.1.1.7 lineage in GISAID.

The VOC strain, as all strains, continues to mutate. Because of its significant replicative advantage, any accrued mutations are given an opportunity to spread (potentially worldwide), depending on their own replicative advantage with respect to the bulk VOC strain, or higher ability to infect seroprevalent individuals. Here, we focus on spike L18F mutation. The first occurrence of this leucine-to-phenylalanine substitution in residue 18 has been reported in a VOC strain genome collected on December 4, 2020 (GISAID ID: EPI_ISL_720875). As of February 5, 2021, as much as 850 spike L18F VOC genomes have been reported in England. Of note, in autumn 2020, that is, before the VOC lineage has become the dominant strain, the spike L18F substitution was an ubiquitous mutation in England. Till February 5, 2021, most of L18F non-VOC genomes in England (97.5%, 25 061 out of 25 712) were found within an A222V strain termed 20A.EU1, that started expanding in England in mid-August 2020 and constituted 63% of genomes sequenced in England in November 2020.^8^ The fraction of spike L18F mutation in the expanding 20A.EU1 strain was slowly increasing from 35% (1332 out of 3799) in September, 43% (5658 out of 13 046) in October, till 52% (8917 out of 17 470) in November 2020, which may suggest that this mutation was beneficial for growth of the 20A.EU1 strain. Here, we demonstrate the nearly exponential growth of the spike L18F VOC substrain with respect to L18 VOC strains in the 5-week period of December 7, 2020–January 17, 2021, suggesting its high replicative advantage within the population of England.

## Results

In Figure 2 we show the exponential growth of the L18F VOC substrain in England in the five-week period of December 7, 2020–January 17, 2021, in relation to the VOC genomes non-mutated at residue 18, denoted L18. In the considered period, the ratio of L18F to L18 genomes increased with the fitted growth rate of *q*_L_ = 1.73. If we assume that the L18F VOC and the L18 VOC strains have the same average serial interval of 6.73 days, this value of *q*_L_ gives 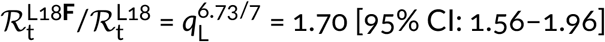. This confidence interval is calculated assuming binomial distribution of the number of the L18F VOC genomes in each week (see Methods). The confidence interval calculated from 1.96 × standard error of the slope, [1.61–1.79], is narrower, which means that nearly perfect co-linearity of five data points is somewhat coincidental, and the binomial distribution-based CI is the proper estimate. This analysis, based on currently very incomplete data, suggests high replication advantage of the L18F VOC substrain in relation to the remaining VOC genomes. This finding is supported by data from Wales, UK, where the L18F VOC genomes constituted 13% (145 out of 1079) of all VOC genomes reported in January, substantially more than in the same period in England, 3% (760 out of 26 174). The number of genomes in Wales is however too small to perform an analysis analogous to that in Figure 2.

**Figure 2:**
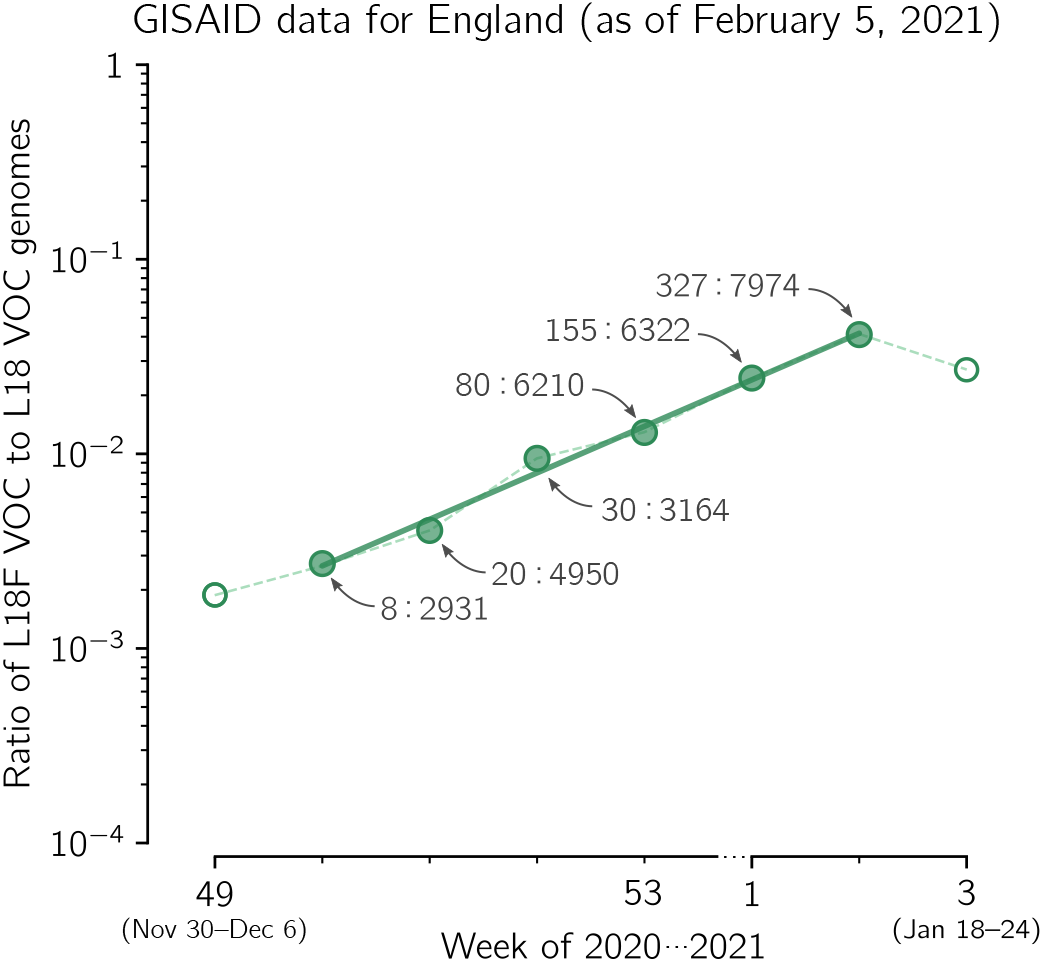
The replicative advantage of the spike L18F substrain in relation to the L18 VOC strains. The ratio of the number o VOC genomes conferring spike L18F mutation to the number of non-mutated (L18) VOC genomes collected in the period of week 49 of 2020–week 3 of 2021 in England. Data aggregated into weeks indicated that *r*_L_ changes from 8 : 2931 = 0.0027 in week 50 of 2020 to 327 : 7974 = 0.041 in the second week of 2021, i.e., 15.0 times, meaning that *r*_L_ increases 15.0^1/5^ ≈ 1.72-fold per week. The trend line is fitted to data points in shown as filled circles. The weekly growth rate of the ratio is *q*_L_ = 1.73. The 95% CI calculated as 1.96 × standard error of the slope is [1.64–1.83]; the 95% CI calculated assuming binomial distribution of substrain genomes is [1.58–2.01]. Data points are labeled with ratios of the counts of both types of genome sequences. The figure is based on GISAID data submitted up till February 5, 2021. The apparent break of the trend the third week of 2021 is possibly caused by incompleteness of data originating from the Alderley Park Lighthouse Lab that in the fitting period contributed about 4/5 (505 out of 620) VOC L18F genomes reported from England.

Of particular concern are the VOC strain mutations occurring in the receptor binding domain (RBD, residues 333–527), especially mutations in the receptor binding motif (RBM, residues 438–506).^9^ These mutations may potentially lead to immune escape mutants, resulting in reinfection of convalescent individuals and lowering efficacy of current vaccines. Propagation of such mutations is facilitated by high replicative advantage of the VOC strain and potential selection due to the increasing number of convalescent or immunized individuals. The VOC-202012/01 strain spike RBM mutations of special concern are substitutions E484K and S494P.

- **E484K**. A first genome has been collected on December 17, 2020 (GISAID ID: EPI_ISL_ 782148) and there were 27 genomes reported from the UK up till February 5, 2021. The same mutation has occurred in fast expanding South African and Brazilian (Manaus) strains that share with the VOC substitution N501Y and additionally have a mutation of residue 417: either K417N (South African strain 501Y.V2)^10^ or K417T (Manaus strain P.1).^11^ It was suggested that E484K may compromise binding of class 2 neutralizing antibodies, while the A501V mutation interferes with binding of class 1 antibodies.^10^ The P.1 strain led in to the surge of infections in Manaus in December 2020 despite high seroprevalence of the population. A study of blood donors indicated that 76% [95% CI: 67–98%] of the population in Manaus had been infected with SARS-CoV-2 by October 2020.^12^
- **S494P**. A first genome has been collected on November 12, 2020 (GISAID ID: EPI_ISL_ 741039), and there were 369 genomes reported from the UK up till February 5, 2021. In an *in silico* study, this substitution has been found to increase complementarity between the RBD and ACE2.^13^ This mutation has been also characterized as an escape mutation by Koenig *et al*.,^14^ who also distinguished six additional “escape” residues in the RBM: G447, Y449, L452, F490, G496, and five outside the RBM but within the RBD: Y369, S371, T376, F377, K378. Among these residues, until February 5, 2021, substitution F490S (first collected on December 13, 2020, GISAID ID: EPI_ISL_736026) was reported in the highest number of genomes (20 genomes in the UK).

## Discussion

We have shown that the VOC substrain conferring L18F mutation is rapidly growing in the UK. Based on data collected in five-week period of December 7, 2020–January 17, 2021, in England we estimated the replicative advantage of this substrain in relation to the remaining VOC strains as 1.70 [95% CI: 1.56–1.96]. Importantly, L18F mutation has also expanded in the South African strain 501Y.V2 defined by three spike mutations K417N, E484K, N501Y (thus sharing with the VOC strain spike mutation N501Y). Among the 501Y.V2 genomes collected after December 1, 2020, the L18F substrain constitutes 40% genomes (97 out of 243, according to GISAID as of February 5, 2021). In Brazil, in strain P.1 defined by three spike mutations K417T, E484K, N501Y (differing from the South African strain 501Y.V2 by substitution K417T instead of K417N) mutation L18F has been found in 92% of genomes (56 out of 61) collected after December 1, 2020. This data suggests replicative advantage of L18F substrains within the VOC, 501Y.V2, and P.1 strains, in, respectively, England, South Africa, and Brazil.

Our finding must be considered with caution until the mechanism promoting faster spread of strains containing L18F substitution is elucidated. Leucine 18 lies in the N-terminal domain (NTD), that has not been typically considered as a target for neutralizing antibodies (Abs). However, there is a growing number of studies showing that the NTD is targeted by Abs and that NTD deletion 69H–70V (characterizing the VOC strain) compromises binding of Abs^15,^[15]. With respect to L18F, an *in vitro* study by Cele *et al*. shows that an African variant L18F, D80A, D215G, K417N, E484K, N501Y, D614G, A701V propagates much faster than a variant without L18F mutation in the presence of plasma antibodies collected from donors infected in the first wave of epidemic in South Africa (June–August, 2020).^16^ Correspondingly, McCallum *et al*. showed that L18F substitution compromises binding of neutralizing Abs.^17^ Findings by Cele *et al*. and McCallum *et al*., together with the increase of L18F variants in 501Y.V2, P.1, and VOC strains, suggests that the replicative advantage of L18F mutants can be partly associated with their ability to infect seroprevalent individuals. In turn, propagation of mutations in escape residues (L18, E484, F490S, or S494) on the VOC strain suggests an increasing selection pressure resulting from the growth of the seroprevalent fraction of the population of England. This trend can be enhanced by the ongoing English vaccination program, in which the relatively large time span between the first and second dose can be a contributing factor.

## Methods

### Estimation of confidence interval for the growth rate of the ratio of strains

To estimate the confidence interval for *k* subsequent weeks *a priori* we assume that genomes of two compared strands in each week of the considered period follow a respective binomial distribution having success probability *p*_i_ = *n*_strain1_/(*n*_strain1_ + *n*_strain2_), where *n*_strain1_ and *n*_strain2_ are the numbers of compared two strain genomes, VOC and non-VOC (in Figure 1) and L18F VOC and L18 VOC (in Figure 2). By sampling from *k* such binomial distributions for the considered time window of *k* weeks 10^5^ times, we obtained 10^5^ series of *k* genome proportions. We performed fitting to each such series to obtain 10^5^ estimates of weekly growth rate of the genomes ratio.

## Data Availability

All data used in this study is available at GISAID initiative webpage https://www.gisaid.org/ upon registration.

## Acknowledgments

This study was supported by the Norwegian Financial Mechanism GRIEG-1 grant 2019/34/H/NZ6/00699 (operated by the National Science Centre Poland). The funding agency had no role in study design, data collection, and analysis, decision to publish, or preparation of the manuscript. We are grateful to Grzegorz Preibisch and Stanisław Giziński for discussions.

## Author contributions

Conceptualization, Frederic Grabowski, Marek Kochańczyk and Tomasz Lipniacki; Data curation and Software, Frederic Grabowski; Investigation, Frederic Grabowski and Marek Kochańczyk; Supervision, Marek Kochańczyk and Tomasz Lipniacki; Visualization, Frederic Grabowski; Writing – original draft, Tomasz Lipniacki; Writing – review & editing, Marek Kochańczyk.

## Corresponding authors

Correspondence to Tomasz Lipniacki.

## Competing interests

The authors have no competing interests.

## Data availability

We analyzed publicly available genome datasets retrieved from GISAID, https://www.gisaid.org.

## Notes

### Competing Interest Statement

The authors have declared no competing interest.

### Funding Statement

This study was supported by the Norwegian Financial Mechanism GRIEG-1 grant number 2019/34/H/NZ6/00699 (operated by National Science Centre Poland).

### Author Declarations

This theoretical study is based on data published by GISAID

